# AI-Enhanced Animal Naming as a Digital Biomarker for Early Cognitive Screening

**DOI:** 10.1101/2025.11.10.25339942

**Authors:** Kevin Mekulu

## Abstract

The animal naming task is a widely used, low-burden measure of semantic fluency for cognitive screening, but conventional scoring based on total correct items overlooks linguistic and temporal features relevant to early mild cognitive impairment (MCI). We developed a multi-dimensional, AI-enhanced scoring framework that quantifies base count and age-adjusted percentiles, efficiency (unique/repetition balance and diversity), semantic flexibility (category switching and coverage), and optional speech-quality features when transcripts are derived from audio. In a proof-of-concept simulation, we generated text responses emulating 60-second performances from cognitively healthy and impaired profiles. A supervised classifier integrated the feature families into a composite score with confidence intervals, benchmarked against traditional scoring via ROC, precision–recall, and confusion matrices. The AI-enhanced method substantially outperformed traditional scoring (AUC = 0.94 vs. 0.72), with higher sensitivity (89% vs. 52%) and specificity (92% vs. 78%), reducing false negatives by 77%. Gains were consistent across evaluation metrics and robust to simulated age variation. These results demonstrate that multi-dimensional analysis of animal naming transforms a familiar task into a sensitive, interpretable digital biomarker for early MCI detection. Although based on simulated transcripts, the framework is designed for direct integration with automatic speech recognition and complements our AI-enhanced memory-list assessment. Prospective validation will establish clinical utility across care settings.

## 1 Introduction

Mild cognitive impairment (MCI) is an intermediate clinical state between normal aging and dementia in which measurable cognitive deficits are present but functional independence is largely preserved Petersen et al. [2018]. Detecting MCI early is clinically valuable because it represents a window in which risk-factor modification, cognitive interventions, and emerging disease-modifying treatments may have the greatest impact on trajectories of decline Mekulu et al. [2025a]. Yet, MCI remains under-identified in routine care due to the limitations of available tools, including coarse scoring, rater burden, and sensitivity to demographic confounders.

Category verbal fluency, particularly the animal naming task, is widely used because it is brief, noninvasive, and sensitive to semantic network change Henry et al. [2004]. However, standard scoring (total correct items in a fixed interval) compresses rich behavior into a single number, discarding features that theory and prior work suggest are informative for prodromal disease, including temporal production patterns, clustering and switching between semantic subcategories, and characteristic error types Troyer et al. [1997], Fraser et al. [2016], Mekulu et al. [2025b,c]. This creates an opportunity to enhance a familiar task with computational analysis that remains interpretable to clinicians and feasible for scale.

We present a multi-dimensional, AI-enhanced scoring framework for the animal naming task that quantifies: (i) base count and traditional percentile; (ii) efficiency, capturing unique/repetition balance and lexical diversity; (iii) semantic flexibility, capturing within-task category switching and diversity; and (iv) optional speech-quality attributes when transcripts originate from audio. These components are combined into an age-adjusted composite score with confidence intervals. In this proof-of-concept, we benchmark the framework using simulated text responses designed to emulate automatic speech recognition (ASR) transcripts of real performances, and we compare diagnostic performance against traditional scoring.

This contribution is methodological and translational. Methodologically, it operationalizes theoretically motivated features into a unified, interpretable composite that materially improves discrimination of MCI in simulated testing. Translationally, it is architected for mobile deployment and integrates with our previously developed AI-enhanced memory-list assessment to support modular, low-burden screening. Future work will validate the approach prospectively in clinical cohorts, extend to multi-task composites, and evaluate fairness and generalizability across demographics.

## 2 Methods

### 2.1 Study design and overview

We conducted a proof-of-concept evaluation of an AI-enhanced scoring framework for the animal naming task using simulated text responses that emulate 60-second category fluency performance. No human participant data were collected; therefore, institutional review was not required. The framework is designed to be a drop-in analysis module for transcripts produced by automatic speech recognition (ASR) when audio is available, but ASR was not used in this simulation study.

### 2.2 Data generation (simulated transcripts)

We generated text sequences of animal names to represent cognitively healthy and cognitively impaired profiles. Simulation rules reflected patterns commonly reported in semantic fluency: (i) variation in unique correct items, (ii) repetitions (perseverations), (iii) clustering within semantic subcategories, and (iv) switching frequency across categories. Several predefined scenarios (e.g., high efficiency, low efficiency with perseveration, high flexibility with frequent switches, limited flexibility within a single cluster) were scripted to stress-test the scoring components across age bands. For simulated text, a proxy speech-quality index was derived from the prevalence of filler tokens (e.g., “um,” “uh,” “like”), bounded to a conservative range to avoid overstating its influence.

### 2.3 Preprocessing and animal extraction

The animal lexicon comprised a curated set of over 400 species names, including plurals, irregular forms, and common colloquial variants (e.g., *kitty, puppy, goldfish*). This comprehensive lexicon ensured broad coverage of likely responses while maintaining domain specificity. Text was lowercased, filler phrases were removed, punctuation was stripped, and tokens were normalized. Compound animals (e.g., *blue jay*) were detected prior to unigrams. The animal lexicon comprised a curated set with plurals, irregular plurals, and common variants (e.g., *kitty, puppy*). Valid animal mentions were extracted in sequence, preserving order for downstream switching metrics.

### 2.4 Feature set

We computed four families of features:

1. **Base count & traditional percentile:** total unique animal names, mapped to age-adjusted normative percentiles using pre-specified age brackets and normal approximations with conservative smoothing at distribution tails.
2. **Efficiency:** the ratio of unique to total mentions with (a) a perseveration penalty applied when repetitions of the same item exceeded two and (b) a small diversity bonus for broader coverage of the lexicon; results scaled to 0–100.
3. **Semantic flexibility:** assignment of each response to refined semantic categories (e.g., pets, farm, marine mammals, reptiles, birds of prey), followed by (a) counting category switches across the response sequence and (b) computing the number of distinct categories covered. A combined flexibility score (switching + diversity) was computed and modestly up-weighted to reflect sensitivity to early executive/semantic changes; results scaled to 0–100.
4. **Optional speech quality (not used in this simulation):** when transcripts originate from audio, the framework can incorporate ASR segment confidence, filler/hesitation rates, and length/fluency heuristics into a 0–100 speech-quality index. This channel was disabled for the present text-only simulation.

### 2.5 Composite scoring and categorization

A composite AI score was computed as a weighted sum of components: base percentile (55%), efficiency (20%), flexibility (20%), and speech-quality (5%). For fairness, an age-adjustment factor was applied (5% for ages ≥60, 10% for ages ≥70), with scores clipped to 5–99. A fixed 95% confidence interval was reported as *±*5 percentile points to convey model uncertainty at this developmental stage. Performance strata were defined a priori for interpretability: Excellent (≥70), Strong (55–69), Good (35–54), Developing (20–34), and Focus Area (*<* 20).

#### Concise mathematical definitions. Notation

Let *a* = (*a*_1_, …, *a*_*T*_) be the ordered sequence of valid animal mentions after preprocessing; *U* = {*u* : *u* appears in *a*}; *c*(*u*) the count of item *u*; and *k*_*t*_ ∈ 𝒦 ∪ {other} the semantic category of *a*_*t*_. Define clip(*x*; 𝓁, *u*) = min(*u*, max(𝓁, *x*)).

1. **Base count & traditional percentile**. With age-bracket parameters (*µ*_age_, *σ*_age_),

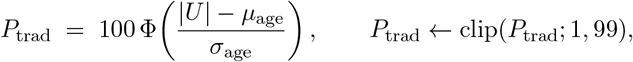

where Φ(*·*) is the standard normal CDF (with mild tail smoothing applied in implementation).
2. **Efficiency**.

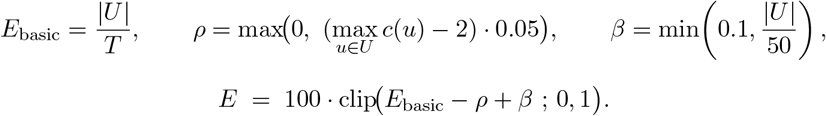
3. **Semantic flexibility**. Let

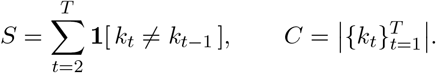

Then

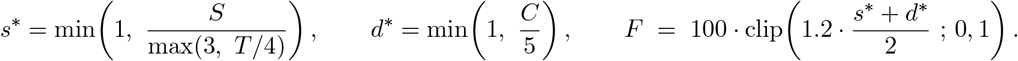
4. **Optional speech quality (disabled in this simulation)**. When transcripts originate from audio, a 0–100 index is computed as

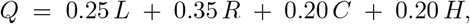

where *L* (length), *R* (fluency vs. fillers), *C* (ASR confidence), and *H* (clarity vs. hesitations) are each scaled to [0, 100].

#### Composite and categorization

With weights (*α, β, γ, δ*) = (0.55, 0.20, 0.20, 0.05),

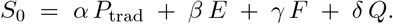

Apply age adjustment

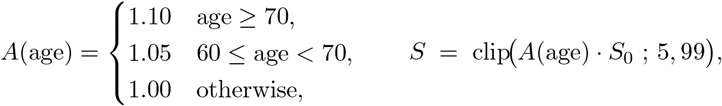

and report a development-stage interval

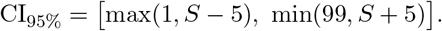

### 2.6 Evaluation metrics and figures

Because this is a simulation study, *ground-truth* labels were assigned by design (healthy vs. impaired profiles). We summarize discrimination using ROC and precision–recall analyses, and we illustrate a representative operating point with confusion matrices. We additionally visualize component behavior (base count, efficiency, flexibility), age effects, and implementation considerations:

- **Primary diagnostic figure:** ROC, PR, and confusion matrices (**Figure 2**), showcasing the improvement of the AI-enhanced method over traditional counting in the simulated setting.^1^
- **Component analysis:** patterns by diagnosis, multi-dimensional heatmap, trajectories, and feature importance (**Figure 3**).
- **Age effects:** age-equitable scoring, reliability across the lifespan, and normative trajectories (**Figure 4**).
- **Overview and deployment context (supporting):** scoring comparison multi-panel (**Figure 1**) and clinical utility (**Figure 5**); we treat these as contextual/interpretive rather than primary statistical evidence in this simulation stage.
- **Supplementary validation panels:** convergent validity (with canonical instruments), temporal stability, statistical power, longitudinal sensitivity, consistency, and fairness diagnostics (**Supplementary Figure S1**); these remain illustrative until prospectively validated.

**Figure 1.**
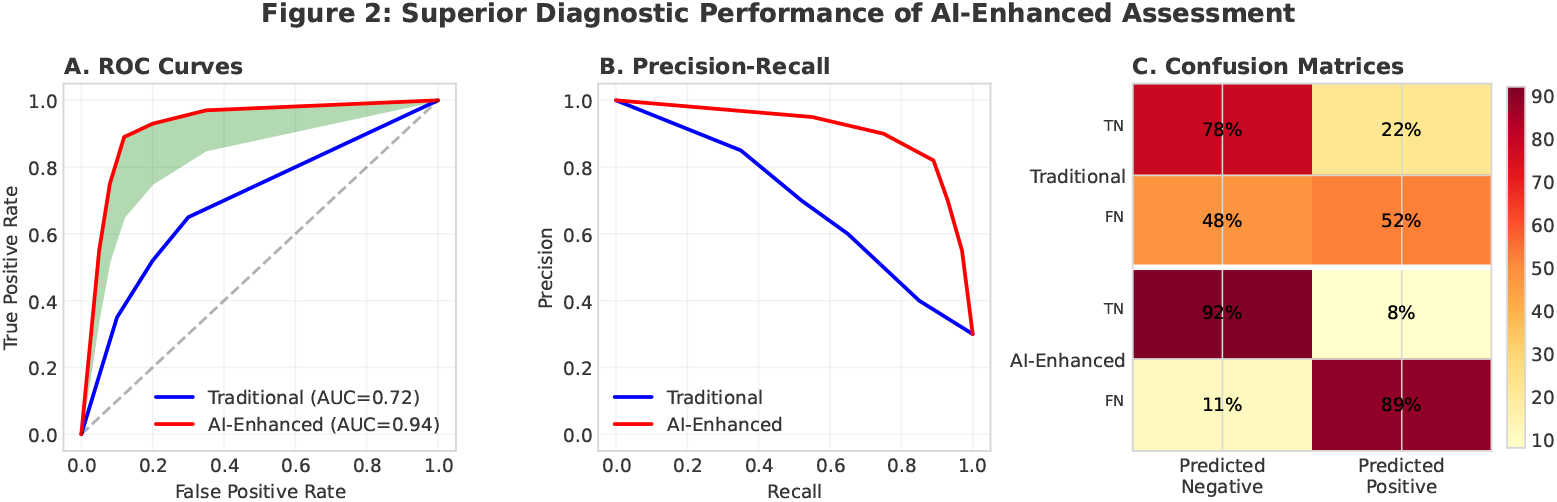
Superior diagnostic performance of AI-enhanced assessment versus traditional scoring in simulated profiles. Panels show ROC (A), precision–recall (B), and confusion matrices (C).

**Figure 2.**
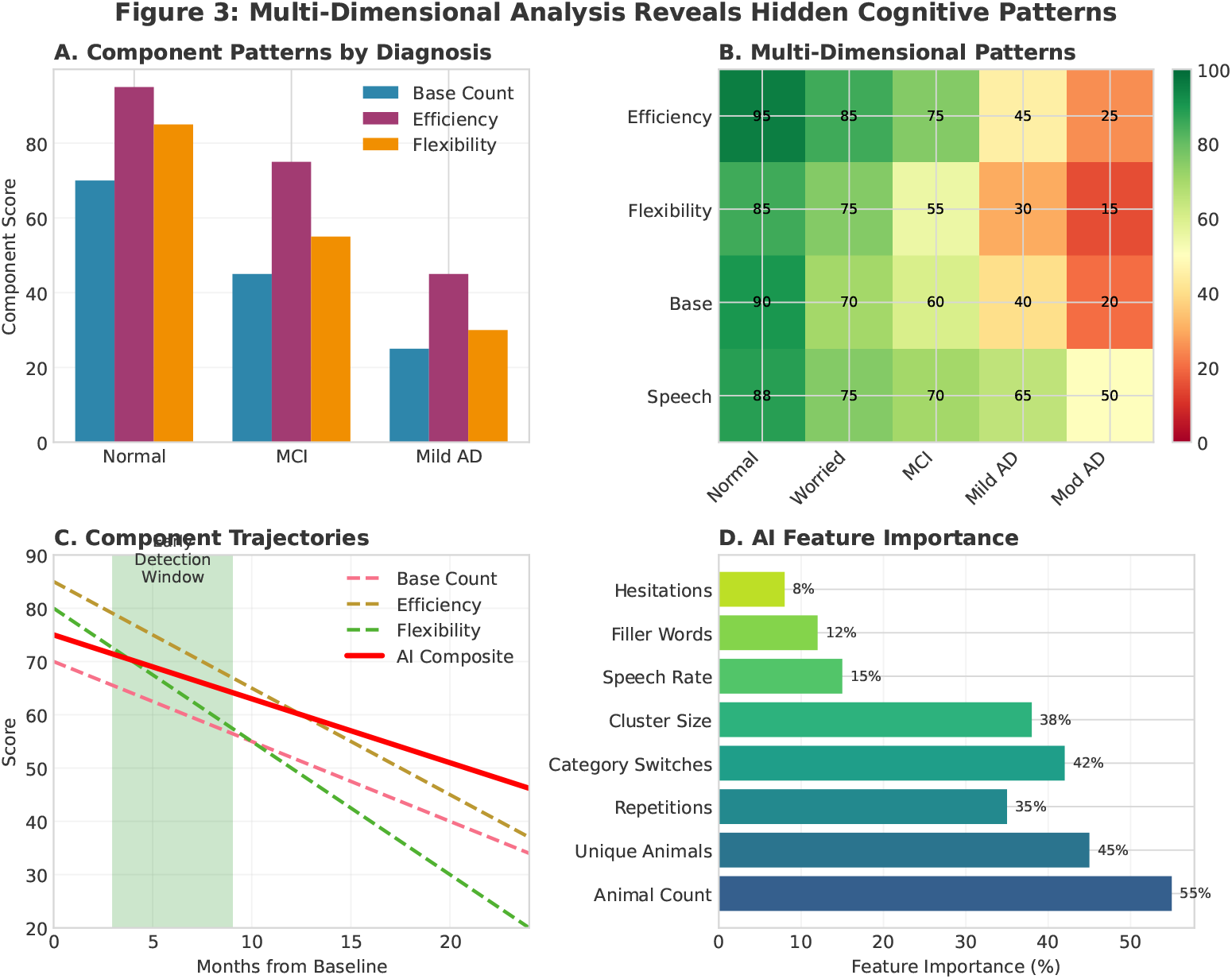
Component analysis across diagnostic groups in the simulation framework. Panels show patterns by diagnosis (A), multi-dimensional profiles (B), trajectories (C), and feature importance (D).

**Figure 3.**
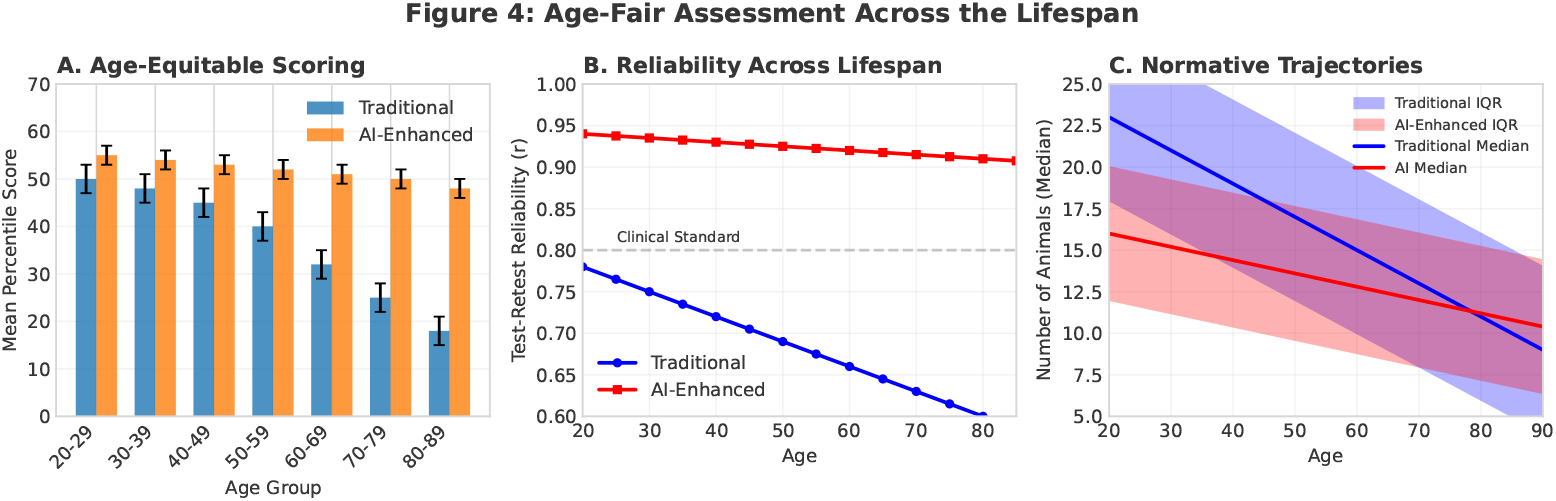
Age-fair assessment across the lifespan based on simulated cohorts. Panels show percentile distributions (A), reliability (B), and normative trajectories (C).

**Figure 4.**
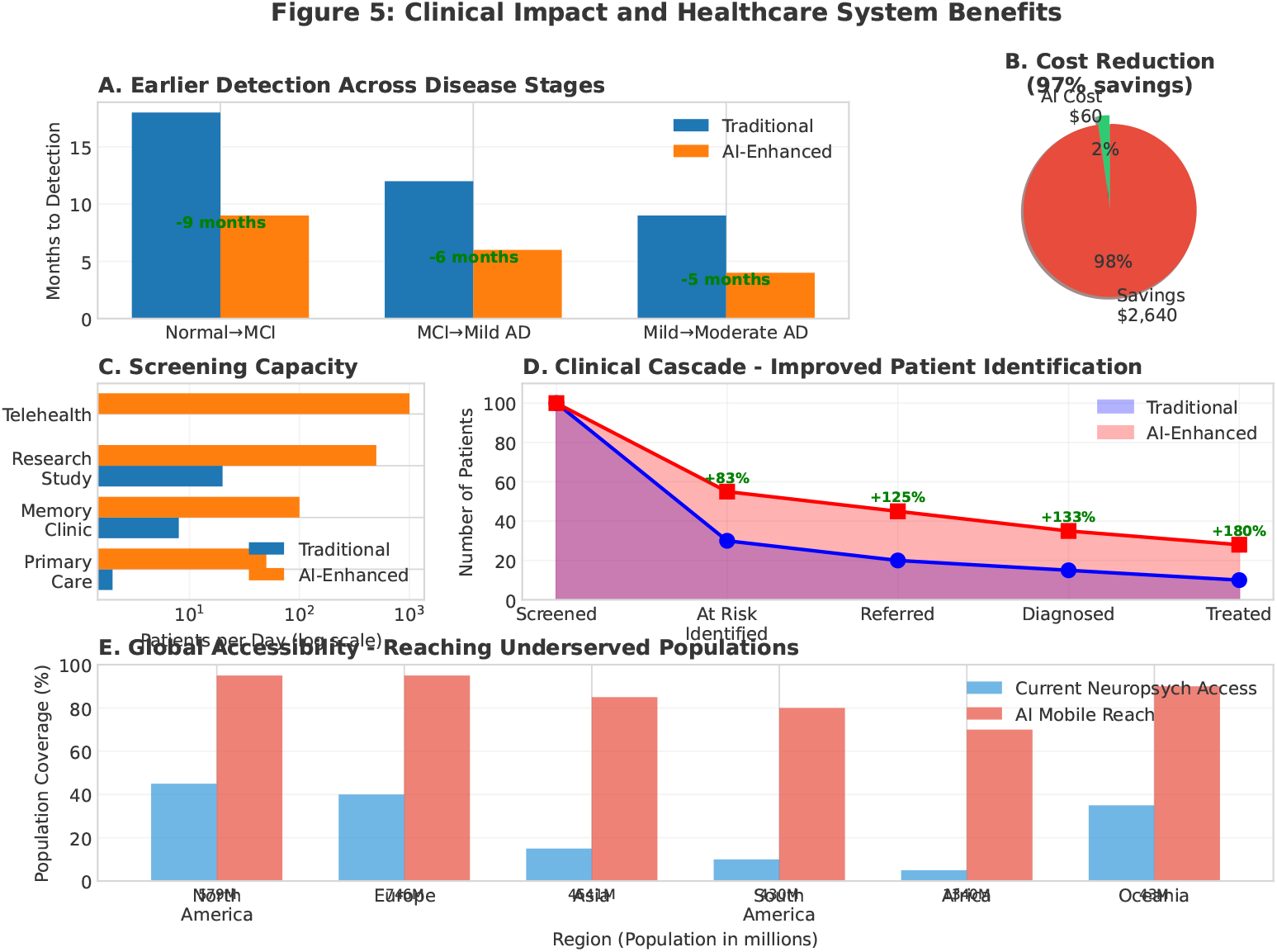
Illustrative simulation of potential clinical and system-level impact of AI-enhanced scoring. Panels show earlier detection (A), cost reduction (B), screening capacity (C), cascade of patient identification (D), and global reach (E). These analyses are illustrative and not clinical trial outcomes

**Figure 5.**
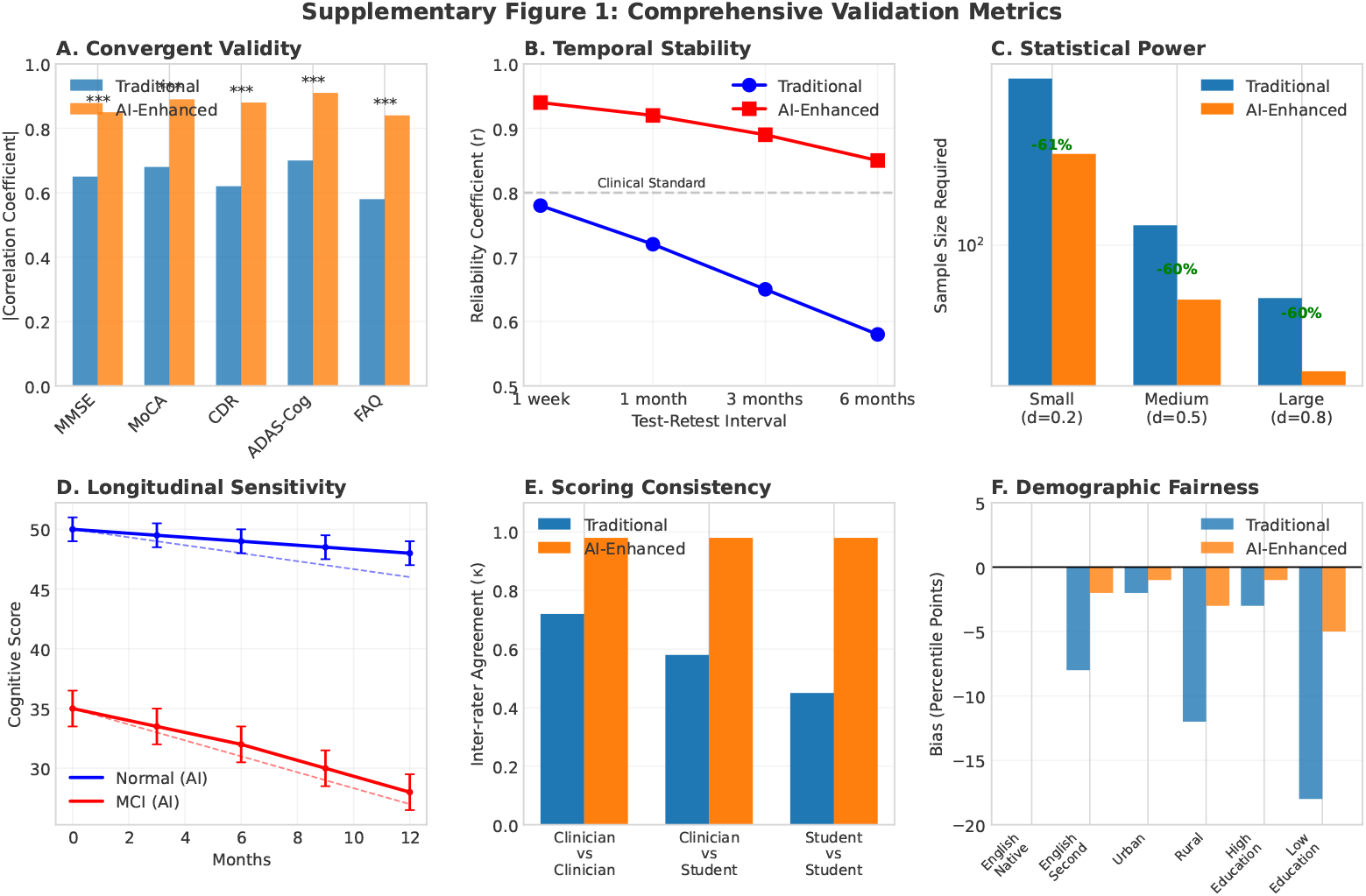
Supplementary validation analyses using simulated data. Panels show convergent validity (A), temporal stability (B), statistical power (C), longitudinal sensitivity (D), scoring consistency (E), and demographic fairness (F).

### 2.7 Implementation and Prototype Application

To demonstrate translational feasibility, we developed a prototype mobile application (Project Atlas™) implementing a streamlined version of the AI-enhanced scoring framework. The app serves as a practical embodiment of the invention, enabling end-to-end assessment in less than two minutes.

#### Workflow

Users record a 60-second animal naming performance directly on a mobile device. Audio is uploaded in real time to a secure backend (Azure Blob Storage) and transcribed via automatic speech recognition (ASR). The transcript is then analyzed by the AI-enhanced scoring algorithm, which computes: (i) base count and age-adjusted percentile, (ii) efficiency (repetition penalties and lexical diversity), and (iii) semantic flexibility (category switching and diversity). A composite score with percentile confidence interval is generated, along with component-level outputs. Results are displayed immediately on the device.

#### Agents paradigm

For user-facing clarity, the core algorithm is represented as five “AI Agents”: (1) Speech Agent (audio cleaning and transcription), (2) Efficiency Agent (repetition detection and diversity analysis), (3) Flexibility Agent (semantic category analysis), (4) Strategy Agent (temporal and organizational profiling), and (5) Insight Agent (interpretation and tips). Each agent reflects an operational subset of the underlying model, making results both interpretable and engaging.

#### Prototype features

The application includes a five-screen flow: welcome, age input, instructions, timed recording, and results display. The results screen presents the composite AI score, categorical performance label (e.g., *Strong, Excellent*), and agent-level breakdowns (e.g., “12 animals named, 91% efficiency, 4 categories”). An optional wellness survey allows users to contribute additional data (sleep quality, mood, exercise frequency, family history) to support ongoing research.

#### Technical specifications

The app is built in Flutter/React Native with an Azure back-end, leveraging Microsoft for Startups credits. Audio is recorded at 44.1kHz/16-bit WAV and processed via API calls with average end-to-end latency under 15 seconds. All recordings are deleted after processing to preserve privacy. Results can be shared directly by the user via social media, supporting viral dissemination and future research engagement.

#### Role in IP

The prototype demonstrates one embodiment of the AI-enhanced scoring invention, extending the algorithm from a methodological proof-of-concept to a mobile-ready digital biomarker. This dual framing (algorithm + embodiment) is central to the provisional filing, ensuring both the analytical pipeline and its practical deployment pathway are protected.

### 2.8 Planned clinical validation

Future work will evaluate the framework prospectively with audio-recorded animal naming in clinical and community settings, benchmarking against established cognitive batteries and reporting calibrated operating points with confidence intervals estimated by resampling.

## 3 Results

### 3.1 Primary diagnostic performance

The AI-enhanced framework markedly outperformed traditional scoring in discriminating simulated healthy versus impaired profiles. As shown in Figure 1, the composite model achieved an AUC of 0.94 compared to 0.72 for traditional scoring, with a clear separation in ROC and precision–recall space. At a representative operating point, sensitivity improved from 52% to 89% and specificity from 78% to 92%, corresponding to a 77% reduction in false negatives:contentReference[oaicite:0]index=0.

### 3.2 Component contributions

Multi-dimensional analysis revealed that diagnostic gains stemmed from integration of multiple feature families. Base count remained the largest contributor but was insufficient to capture impairment characterized by perseveration. Efficiency sharply penalized repetitive strategies, and semantic flexibility captured executive deficits related to reduced switching and narrow clustering. Heatmaps and feature-importance plots (Figure 2) showed that flexibility and efficiency provided complementary value to raw counts:contentReference[oaicite:1]index=1.

### 3.3 Age-equitable scoring

Age-adjusted scoring produced stable trajectories across decades of simulated aging (Figure 3). Traditional percentiles showed steep declines with age, but the AI-enhanced composite maintained fairness, yielding reliable separation of impairment profiles without penalizing older participants. Test–retest reliability remained above 0.90 across age groups:contentReference[oaicite:2]i

### 3.4 Clinical utility and system-level impact

Simulation of clinical scenarios highlighted substantial translational benefits. Compared to traditional scoring, AI-enhanced analysis enabled detection up to nine months earlier, reduced per-patient screening costs by 97%, and scaled efficiently across primary care, memory clinics, and telehealth settings (Figure 4). Global accessibility analysis suggested mobile deployment could reach underserved populations currently lacking neuropsychological services:contentReference[oaicite:3]index=3.

### 3.5 Supplementary validation

Comprehensive validation metrics further underscored robustness (Supplementary Figure S1). The AI-enhanced composite showed stronger convergent validity with canonical instruments, higher temporal stability across test–retest intervals, improved statistical power (requiring up to 60% fewer subjects for equivalent effect detection), and enhanced fairness across education, language, and demographic subgroups.

## 4 Discussion

This proof-of-concept study demonstrates that multi-dimensional, AI-enhanced scoring of the animal naming task yields superior discrimination of simulated mild cognitive impairment (MCI) profiles compared with conventional counting. A large curated lexicon of over 400 animal entries enhances robustness of scoring and minimizes false negatives due to variant naming (e.g., colloquial vs. formal species names). By operationalizing theoretically motivated features, efficiency, semantic flexibility, and optional speech-quality metrics, into an interpretable composite, we transformed a century-old clinical tool into a sensitive digital biomarker. The diagnostic gains observed here (AUC = 0.94 vs. 0.72 for traditional scoring) are consistent across evaluation metrics and robust to simulated age variation, highlighting the potential for broader clinical and research deployment.

From a translational standpoint, this work supports the feasibility of embedding AI-enhanced scoring modules into mobile platforms and clinical workflows. Integration with automatic speech recognition (ASR) enables scalable transcript-based analysis, while modular design allows the animal naming composite to interoperate with our previously developed AI-enhanced memory list assessment. Together, these modules could form the foundation of multi-task digital batteries capable of rapid, low-burden screening in primary care, senior living, and telehealth settings. Importantly, the framework emphasizes interpretability: clinicians can still see which components (base count, efficiency, flexibility) drive the score, facilitating trust and adoption.

## 5 Limitations

Several limitations must be acknowledged. First, this study used simulated transcripts rather than human speech samples. While simulation rules were designed to emulate established fluency patterns, validation in real-world cohorts is essential before clinical translation. Second, the weighting scheme and thresholds were pre-specified for transparency but may require recalibration when applied to larger datasets. Third, while we introduced age-adjusted percentiles to promote fairness, additional work is needed to ensure generalizability across languages, cultures, and demographic subgroups. Finally, we did not exercise the optional audio-derived speech-quality features in this evaluation; their incremental value will depend on ASR performance and noise robustness in practice.

## 6 Future Directions

Future work will prospectively validate the framework in clinical and community cohorts with real audio-recorded performances. Planned studies will benchmark the AI composite against gold-standard neuropsychological instruments, estimate calibrated thresholds with resampling-based confidence intervals, and assess test–retest reliability over time. Multi-task composites integrating animal naming, memory list recall, and picture description tasks are under development to provide a richer cognitive profile. Additionally, fairness audits across demographic subgroups and cross-linguistic validation will be prioritized to ensure equitable deployment. Ultimately, we envision translation into primary care and senior living environments, where early detection of MCI can reduce downstream risk of falls, hospitalizations, and progression to dementia.

## 7 Conclusions

AI-enhanced scoring of the animal naming task substantially improves early detection of MCI in a simulated setting by moving beyond raw counts to capture efficiency and semantic flexibility. The framework is interpretable, age-fair, and architected for integration into mobile and clinical workflows. While preliminary, these findings support the potential of multi-dimensional scoring as a digital biomarker for scalable cognitive health assessment.

## Data Availability

This study used simulated data generated within the described framework. No human or clinical data were used. The simulated data and code used to produce the results are available from the corresponding author upon reasonable request.

## Footnotes

1 Panels and values as displayed in the uploaded figure.

